# Postvaccination SARS-CoV-2 infection among healthcare workers – A Systematic Review and meta-analysis

**DOI:** 10.1101/2021.10.04.21264542

**Authors:** Saurabh Chandan, Shahab R. Khan, Smit Deliwala, Babu P. Mohan, Daryl Ramai, Ojasvini C. Chandan, Antonio Facciorusso

## Abstract

**INTRODUCTION:** Healthcare workers (HCWs) remain on the front line of the battle against SARS-CoV-2 and COVID-19 infection, and are among the highest groups at risk of infection during this raging pandemic. We conducted a systematic review and meta-analysis to assess incidence of postvaccination SARS-CoV-2 infection among vaccinated HCWs.

**METHODS:** We searched multiple databases from inception through August 2021 to identify studies that reported on incidence of postvaccination SARS-CoV-2 infection among HCWs. Meta-analysis was performed to determine pooled proportions of COVID-19 infection in partially and fully vaccinated individuals.

**RESULTS:** Eighteen studies with 228,873 HCWs were included in the final analysis. Total number of partially vaccinated, fully vaccinated, and unvaccinated HCWs were 132,922, 155,673 and 17505, respectively. Overall pooled proportion of COVID-19 infections among partially/fully vaccinated and unvaccinated HCWs was 2.1% (95% CI 1.2-3.5). Among partially vaccinated, fully vaccinated and unvaccinated HCWs, pooled proportion of COVID-19 infections was 3.7% (95% CI 1.8-7.3), 1.3% (95% CI 0.6-2.9), and 10.1% (95% CI 4.5-19.5), respectively.

**DISCUSSION:** Our analysis shows the risk of COVID-19 infection in both partially and fully vaccinated HCWs remains exceedingly low when compared to unvaccinated individuals. There remains an urgent need for all frontline HCWs to be vaccinated against SARS-CoV-2 infection.

## 1. INTRODUCTION

Healthcare workers (HCWs) remain on the front line of the battle against SARS-CoV-2 and COVID-19 infection, and through interactions in the workplace related to care and proximity to patients, in addition to household and community interactions, they are among the highest groups at risk of infection during this raging pandemic.^1^ At the peak of the pandemic, a large systematic review of 594 studies noted that 152 888 COVID-19 infections and 1413 deaths occurred among HCWs globally. The overall infection and death trends among HCWs followed that of the general population.^2^ More recent estimates suggest that more than 233 million cases of the novel coronavirus have been diagnosed globally, resulting in more than 4 million deaths.

In December 2020, two messenger RNA (mRNA) vaccines, the BNT162b2 vaccine from Pfizer–BioNTech and the mRNA-1273 vaccine from Moderna, were approved by the Food and Drug Administration under Emergency Use Authorization for use among persons 16 years of age or older (for the BNT162b2 vaccine) or among those 18 years or older (for the mRNA-1273 vaccine). Recent data suggests that these vaccines are highly effective under real-world conditions in preventing symptomatic Covid-19 in HCWs, including those at risk for severe Covid-19.^3-5^ Despite the global push for vaccination, studies show that vaccine hesitancy among HCWs is still common with acceptance rates ranging widely from 27.7% to 77.3%. Demographic variables such as men, older age and physicians were positive predictive factors, whereas concerns for safety, efficacy and effectiveness and distrust of the government were barriers. ^6^

In the recent months, there have been further concerns about emergence of SARS-CoV-2 variants, including variants first reported in the United Kingdom (B.1.1.7), South Africa (B.1.351), Brazil (P.1), California (B.1.427/B.1.429), and India (B.1.617). ^7, 8^ As vaccine effective data emerges, we conducted a systematic review and meta-analysis to assess incidence of postvaccination SARS-CoV-2 infection among vaccinated HCWs.

## 2. METHODS

### 2.1. Search strategy

The relevant medical literature was searched by two authors (SC, DR) for studies reporting on the incidence and outcomes of postvaccination COVID-19 infection among HCWs. A systematic and detailed search was run in August 2021 in Ovid EBM Reviews, ClinicalTrials.gov, Ovid Embase (1974+), Ovid Medline (1946+ including epub ahead of print, in-process & other non-indexed citations), Scopus (1970+), Web of Science (1975+) and Google Scholar. Literature search was performed to include studies published in all languages, and in the case of non-English studies, electronic language translation service was used to convert the text to English. An example search strategy using EMBASE is presented as Supplementary Appendix-1. Articles were included if data with regards to incidence of postvaccination COVID-19 infection was presented. Only cohort studies were eligible for inclusion. All other study designs including, case series of less than 10 patients, case reports, review articles, and guidelines were excluded.

As the included studies were observational in design, the MOOSE (Meta-analyses Of Observational Studies in Epidemiology) Checklist was followed ^9^ and is provided as Supplementary Appendix-2. PRISMA Flowchart for study selection and PRISMA Checklist ^10^ are provided as Supplementary Figure-1 and Supplementary Appendix-2, respectively. Reference lists of evaluated studies were further examined to identify other studies of interest.

### 2.2. Data abstraction and quality assessment

Data on study-related outcomes from the individual studies were abstracted independently onto a standardized form by at least two authors (SC, SRK). Authors (DR, OCC) cross-verified the collected data for possible errors and two authors (SC, SRK) performed the quality scoring independently. We used the Newcastle-Ottawa scale to assess the quality of cohort studies. ^11^ This quality score consisted of 8 questions, the details of which are provided in Supplementary Table 1.

### 2.4. Outcomes assessed

1. Overall pooled proportion of positive COVID-19 infections in fully, partially, and unvaccinated HCWs
2. Pooled proportion of COVID-19 infections in partially vaccinated HCWs
3. Pooled proportion of COVID-19 infections in fully vaccinated HCWs
4. Pooled proportion of COVID-19 infections in unvaccinated HCWs
5. Pooled proportion of vaccinated HCWs hospitalized for COVID-19 infection
6. Pooled proportion of vaccinated HCWs admitted to ICU for COVID-19 infection
7. Pooled proportion of vaccinated HCWs died from COVID-19 infection

### 2.5. Statistical analysis

We used meta-analysis techniques to calculate the pooled estimates in each case following the methods suggested by DerSimonian and Laird using the random-effects model and results were expressed in terms of pooled proportion (PP) along with relevant 95% confidence intervals (CIs). ^12^ When the incidence of an outcome was zero in a study, a continuity correction of 0.5 was added to the number of incident cases before statistical analysis. ^13^ A p-value of <0.05 was defined as statistically significant. We assessed heterogeneity between study-specific estimates by using Cochran Q statistical test for heterogeneity, 95% confidence interval (CI) and the I^2^ statistics. ^13-15^ In this, values of <30%, 30% - 60%, 61% - 75%, and >75% were suggestive of low, moderate, substantial, and considerable heterogeneity, respectively. We assessed publication bias, qualitatively, by visual inspection of funnel plot and quantitatively, by the Egger test. ^16^ When publication bias was present, further statistics using the fail-Safe N test and Duval and Tweedie’s ‘Trim and Fill’ test was used to ascertain the impact of the bias. ^17^

All analyses were performed using Comprehensive Meta-Analysis (CMA) software, version 3 (BioStat, Englewood, NJ).

## 3. RESULTS

### 3.1. Search results and population characteristics

All search results were exported to Endnote where 22 obvious duplicates were removed leaving 92 citations. Eighteen studies with 228,873 HCWs were included in the final analysis. Total number of partially vaccinated, fully vaccinated, and unvaccinated HCWs were 132,922, 155,673 and 17505, respectively. A schematic diagram demonstrating our study selection is illustrated in Supplementary Figure 1.

### 3.1. Characteristics and quality of included studies

Six studies originated from India,^18-23^ seven from USA, ^1, 24-29^, two from Israel, ^30, 31^ and one each from Pakistan, ^32^ United Kingdom ^33^ and Indonesia. ^34^ Further details of patient characteristics, follow up time and type of infection, symptomatic or asymptomatic is presented in Table(s) 1-2

Ten of the included studies were retrospective in design while four were prospective. Based on the New-Castle Ottawa scoring system, all included studies were considered to be of high quality.

### 3.3. Meta-analysis outcomes

1. **Overall pooled proportion of positive COVID-19 infections in fully, partially, and unvaccinated HCWs -** Across 18 studies, the overall pooled proportion of COVID-19 infections was 2.1% (95% CI 1.2-3.5; I^2^ 99.5%) Figure 1.

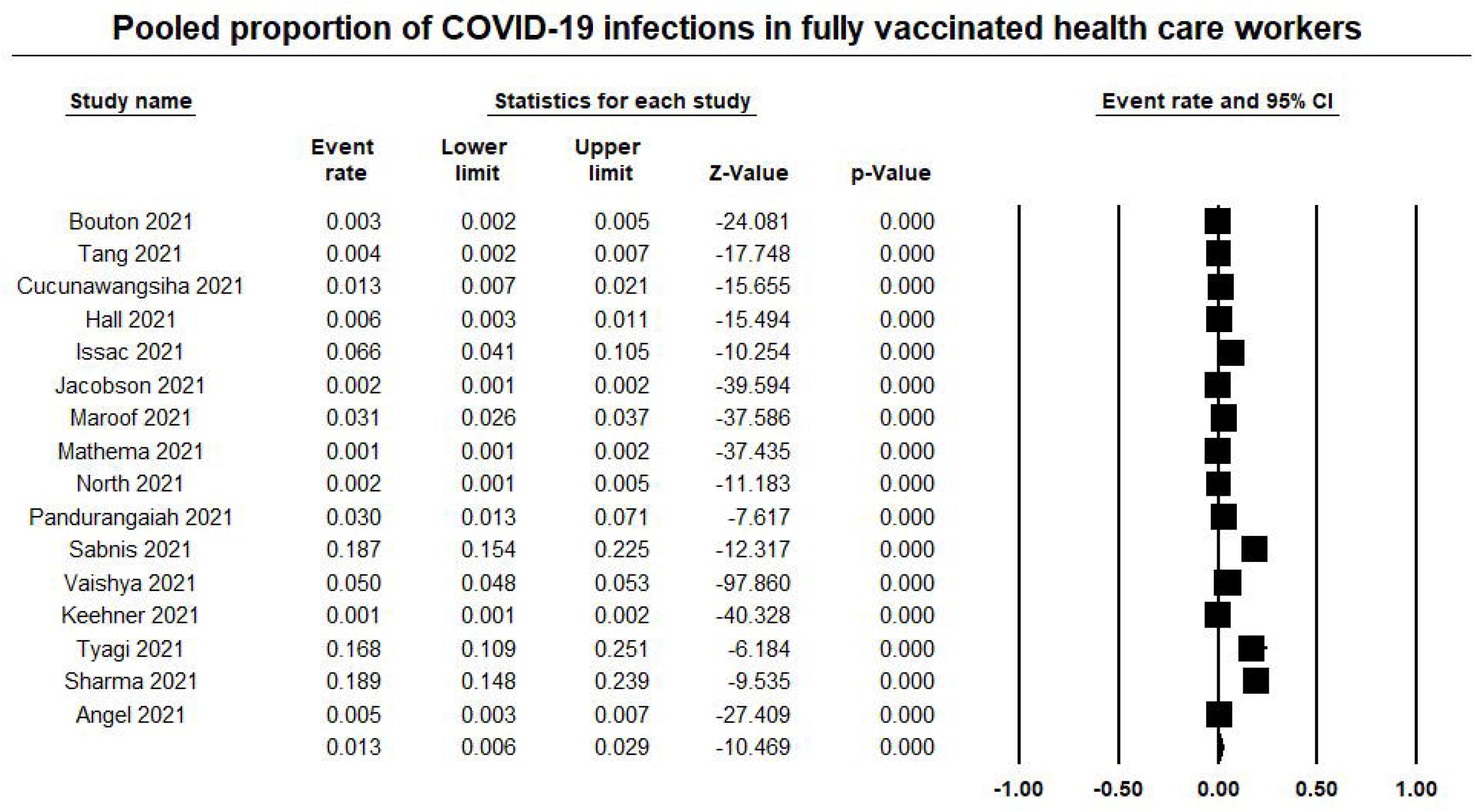
2. **Pooled proportion of COVID-19 infections in partially vaccinated HCWs –** Among partially vaccinated HCWs, across 14 studies, the overall pooled proportion of COVID-19 infections was 3.7% (95% CI 1.8-7.3; I^2^ 99.2%) Figure 2.

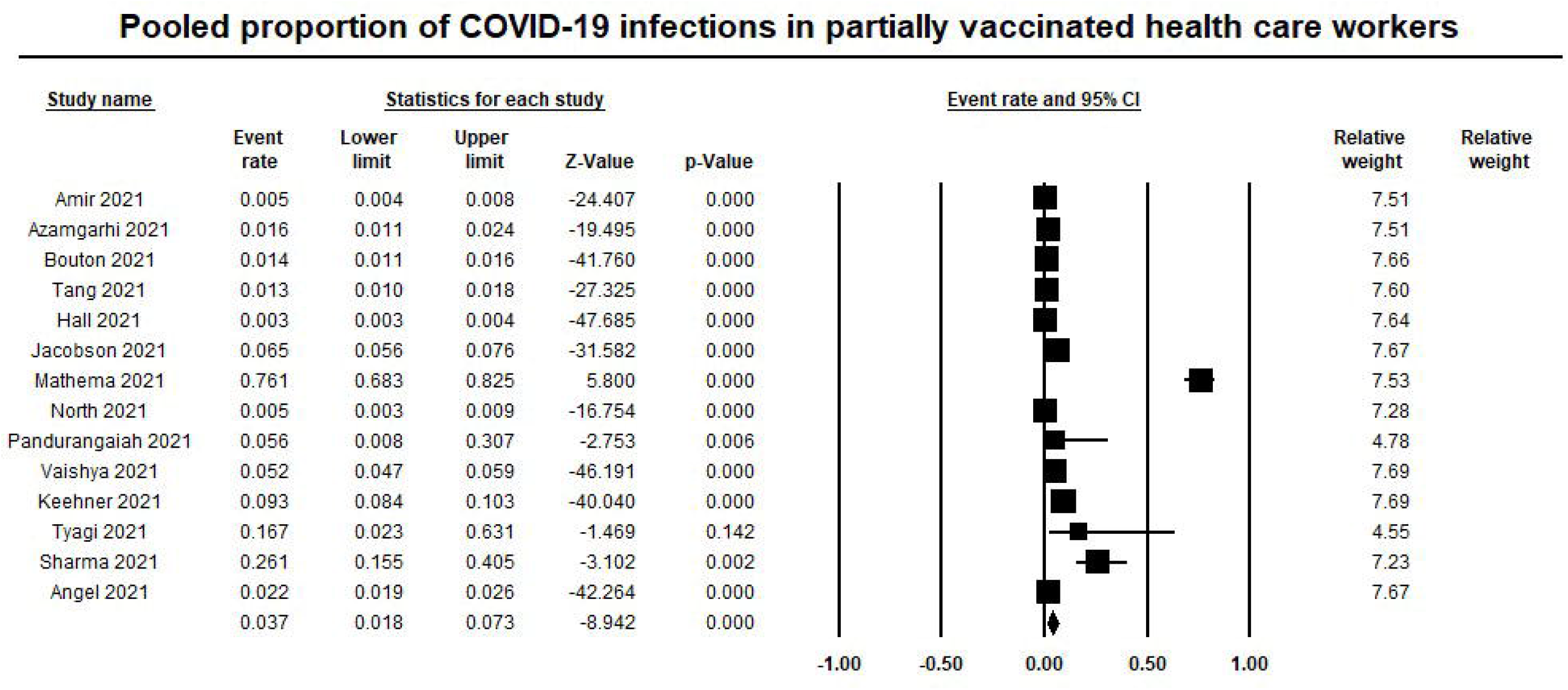
3. **Pooled proportion of COVID-19 infections in fully vaccinated HCWs –** Among fully vaccinated HCWs, across 16 studies, the overall pooled proportion of COVID-19 infections was 1.3% (95% CI 0.6-2.9; I^2^ 99.3%) Figure 3.

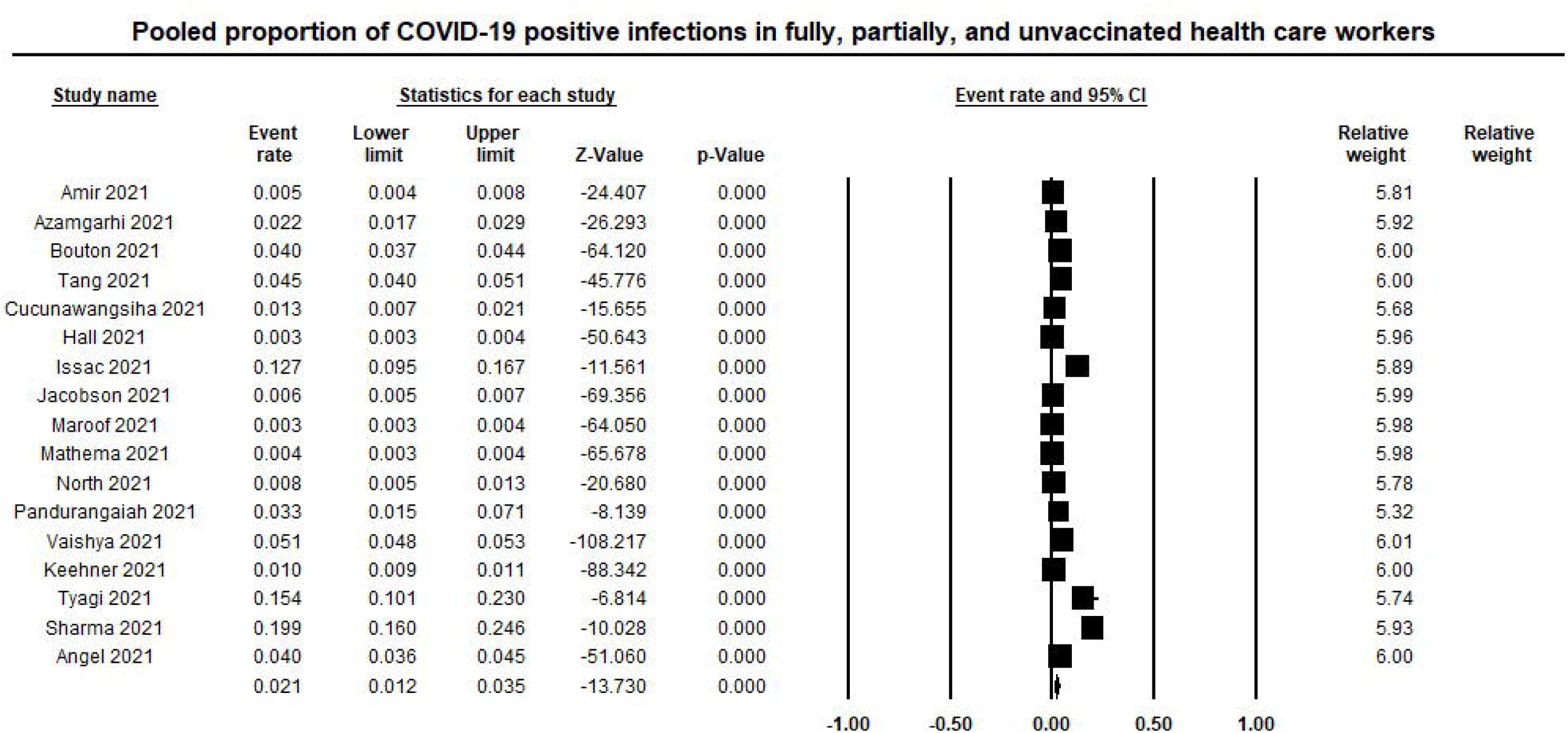
4. **Pooled proportion of COVID-19 infections in unvaccinated HCWs –** Among unvaccinated HCWs, across 8 studies, the overall pooled proportion of COVID-19 infections was 10.1% (95% CI 4.9-19.5; I^2^ 99.5%) Supplementary Figure 2.
5. **Pooled proportion of vaccinated HCWs hospitalized for COVID-19 infection –** The overall pooled proportion of both fully and partially vaccinated HCWs hospitalized for COVID-19 infection was 5.7% (95% CI 3.5-9.1; I^2^ 48.4%) Supplementary Figure 3.
6. **Pooled proportion of vaccinated HCWs admitted to ICU for COVID-19 infection -** The overall pooled proportion of both fully and partially vaccinated HCWs requiring intensive care unit admission for COVID-19 infection was 2.6% (95% CI 0.4-15.4; I^2^ 84%) Supplementary Figure 4.
7. **Pooled proportion of vaccinated HCWs died from COVID-19 infection -** The overall pooled proportion of both fully and partially vaccinated HCWs dying for COVID-19 infection was 1.2% (95% CI 0.3-5.7; I^2^ 72.6%) Supplementary Figure 5.

## 4. Validation of meta-analysis results

### 4.1. Sensitivity analysis

To assess whether any one study had a dominant effect on the meta-analysis, we excluded one study at a time and analyzed its effect on the main summary estimate. We found that exclusion of any single study did not significantly affect the primary outcome or influence the heterogeneity.

### 4.2. Heterogeneity

We assessed dispersion of the calculated rates using the I^2^ percentage values as reported in the meta-analysis outcomes section. We found considerable heterogeneity in our outcomes. This is likely due to variety in the sample size of each individual study, the type of COVID-19 vaccine administered and variation in mean follow up time.

### 4.3. Publication bias

Based on visual inspection of the funnel plot for our study outcomes, we found no evidence of publication bias. Quantitative assessment demonstrated an Egger’s 2-tailed p-value of 0.4 Supplementary Figure 11(a-c).

## 5. DISCUSSION

Our analysis shows the risk of COVID-19 infection in both partially and fully vaccinated HCWs remains exceedingly low when compared to unvaccinated individuals. We found that while the pooled proportion of unvaccinated HCWs contracting COVID-19 was as high as 47%, this decreased to 3.7% for partially vaccinated and 1.3% for fully vaccinated HCWs. At the time of writing, the COVID-19 pandemic continues to rage across the world and HCWs account for a large number of infected people. ^35^ These individuals are both not only victims of the disease, but also potential spreaders. ^36^ Therefore, protecting HCWs from SARS-CoV-2 infection would not only be beneficial for themselves, but also for their household contacts and patients. Vaccine acceptance among HCWs and hesitancy remains a concern with studies showing that nurses and assistant nurses were less prone to accept vaccination against COVID-19 than physicians. ^37^ Our study is crucial in that it is the first in literature to systematically review and analyze the incidence of COVID-19 infections among partially/fully vaccinated or unvaccinated HCWs.

In December 2020, two messenger RNA (mRNA) vaccines, the BNT162b2 vaccine from Pfizer–BioNTech and the mRNA-1273 vaccine from Moderna, were approved by the Food and Drug Administration under Emergency Use Authorization for use among persons 16 years of age or older (for the BNT162b2 vaccine) or among those 18 years or older (for the mRNA-1273 vaccine). ^38, 39^ The U.S. Advisory Committee on Immunization Practices recommended the prioritization of health care personnel during early-phase distribution of these vaccines to ensure that the spread of infection in health care settings was reduced. Vaccination of health care personnel in the United States was initiated in December 2020, and by early March 2021, more than half the frontline health care personnel in the United States had been vaccinated with Covid-19 vaccines. ^40^ Despite this, vaccine hesitancy in the general population and among HCWs remains a concern. ^41, 42^ A recent review by Biswas et al. reported that the prevalence of COVID-19 vaccination hesitancy worldwide in healthcare workers ranged from 4.3 to as high as 72%, with an averagel1of 22.51% across all studies with 76,471 participants. The authors reported concerns about vaccine safety, efficacy, and potential side effects as top reasons for COVID-19 vaccination hesitancy in HCWs. Given the high prevalence of COVID-19 vaccine hesitancy in healthcare workers, communication and education strategies along with mandates for clinical workers should be considered to increase COVID-19 vaccination uptake in these high-risk individuals. ^43^ Studies have also shown that vaccination amongst health care workers is associated with a substantial reduction in COVID-19 cases in household contacts consistent with an effect of vaccination on transmission. ^44^

At the peak of the pandemic, assessing published data between 01 May to 09 July 2020, researchers found that a significant number of HCW were reported to be infected with COVID-19 during the first 6 months of the COVID-19 pandemic, with a prevalence of hospitalization of 15.1% and mortality of 1.5%. 45 With that in mind, we analyzed the pooled prevalence of COVID-19 infections among HCWs who declined vaccinations and those who either received one or both the vaccines. Our analysis shows that only 5.7% of vaccinated HCWs required hospitalization for COVID-19 infection, with 2.6% needing ICU level-of-care. Mortality associated with COVID-19 infection in partially and/or fully vaccinated HCWs remained low at 1.2%.

There are several strengths to our analysis. First, we conducted a systematic literature search with well-defined inclusion criteria, careful exclusion of redundant studies, inclusion of good quality studies with detailed extraction of data and rigorous evaluation of study quality. All the included studies in our analysis were of high quality. Secondaly, our analysis included outcomes separately for partially and fully vaccinated HCWs. We calculated the pooled proportion of HCWs requiring hospitalization, ICU admissions and also assessed the mortality associated with COVID-19 infection in vaccinated HCWs. Finally, our analysis included studies from different geographical locations, making our results more generalizable and clinically relevant. However, there are also several limitations to this study, most of which are inherent to any meta-analysis. Firstly, at the time of writing and based on our literature search, a total of 18 studies were included in our analysis. As active research continues to be conducted on COVID-19 pandemic, it is possible that we may not have captured all the literature, especially studies not indexed in major databases and/or studies which are published-ahead-of-print. Secondly, we were unable to sub-group our outcomes based on which particular vaccine was administered to the cohort of HCWs. Thirdly, we were unable to determine the mean time to infection occurrence post vaccination, as this information was not consistently reported in the all the studies. There was considerable heterogeneity in our study outcomes likely due to variation in the type of vaccination used and median time to infection occurrence. Lastly, ten of the eighteen studies included in our analysis were retrospective in design which may have resulted in selection bias.

Nevertheless, our study is the first in literature to assess the pooled incidence of postvaccination SARS-CoV-2 infection among health care workers around the world. Our results show decreased incidence of COVID-19 infection as well as decreased incidence of hospitalization, ICU admission and deaths, amongst vaccinated HCWs. Our findings support the urgent need for HCWs to consider getting vaccinated against COVID-19.

## Data Availability

N/A

