## Supplementary material for "Postvaccination SARS-CoV-2 infection among healthcare workers – A Systematic Review and meta-analysis": Table 2

**Table 2 – Study Details**

| Author | Asymptomatic COVID-19 Positive | | | Symptomatic COVID-19 Positive | | | Time to symptom onset / Median F/u time | COVID-19 Outcomes | | |
| --- | --- | --- | --- | --- | --- | --- | --- | --- | --- | --- |
|  | Partial Vaccination | Full Vaccination | Unvaccinated | Partial Vaccination | Full Vaccination | Unvaccinated |  | Hospitalized | ICU | Death |
| Amit, 2021 | 11/ 22 | -- | -- | 11/ 22 | -- | -- | 3.5 d (0–10 d) [Median] | -- | -- | -- |
| Azamgarhi, 2021 | 2/ 23 | -- | 6/ 26 | 21/23 | -- | 20/26 | 3.5 d (range 0–10) | -- | -- | -- |
| Bouton, 2021 | 20/96 | -- | -- | 76/96 | -- | -- | 1-14 d (67), 15+ d (29) | -- | -- | -- |
| Tang, 2021 | 29/51 | | 79/185 | 22/51 | | 106/185 | Median F/U time - 81 d (unvaccinated); 72 d (vaccinated) | -- | -- | -- |
| Cucunawangsiha, 2021 | -- | 2/13 | -- | -- | 11/13 | -- | 5 (range 2 - 11) | -- | -- | -- |
| Hall, 2021 | 15/80 | | 676/977 | 65/80 | | 543/977 | Median F/U time - 59 days post-first dose (median 21, IQR 13–31), 39 days post- second dose (23, 17–28) | -- | -- | -- |
| Issac, 2021 | -- | -- | -- | -- | 16/243 | -- | 65 (20 - 91) | 0/16 (FV) | 0/16 (FV) | 0/16 (FV) |
| Jacobson, 2021 | -- | 8/39 | -- | -- | 31/39 | -- | 8 days (IQR, 3–18) | 2/189 (FV+PV) | -- | 0/189 (FV+PV) |
| Maroof, 2021 | -- | 117/124 [Asymptomatic/Mild] | -- | -- | 7/124 | -- | 36.5 d (IQR, 26-62) [Full Vaccination] | -- | -- | 0/124 (FV) |
| Mathema, 2021 | 21/138 | | -- | 117/138 | -- | -- | 105/138 (1-113 d) [Partial Vaccination], 33/138 (4-104 d) [Full Vaccination] | 5/138 (FV+PV) | -- | -- |
| North, 2021 | 4/10 | 2/3 | 1/6 | -- | -- | -- | Full vaccination - 17 days [12, 22], Partial vaccination - 26 days [15, 36] | 0/13 (FV+PV) | -- | -- |
| Pandurangaiah, 2021 | 2/182 | | | -- | -- | -- | NR | 1/ 6 (FV+PV) | 0/6 (FV+PV) | 0/6 (FV+PV) |
| Sabnis, 2021 | -- | -- | -- | -- | 86/461 | -- | 38.42±12.2 days (range 17-70 days) | 10/86 (FV) | 2/86 (FV) | 1/86 (FV) |
| Vaishya, 2021 | -- | -- | -- | -- | -- | -- | < or >2 weeks | 83/1438 (FV + PV) | 3/1/1438 (FV + PV) | 0/1438 (FV + PV) |
| Keehner, 2021 | -- | -- | -- | -- | -- | -- | Dose 1 - 1-7d (145), 8-14d (125), 15-21d (47), d22 (15); Dose 2 - 1-7d (22), 8-14d (8), 15d or later (7) | -- | -- | -- |
| Tyagi, 2021 | -- | -- | -- | 1/19 | 18/19 | -- | 34.8 d (2-51 d) [Full Vaccination] | 1/ 19 (FV + PV) | -- | -- |
| Sharma, 2021 | -- | -- | -- | 59 (Mild), 6 (Moderate) | | -- | 46 (28.2, 54.7) d [Full Vaccination] | -- | -- | -- |
| Angel, 2021 | 63/ 5761 | 19/5372 | 31/757 | 64/ 5761 | 8/5372 | 85/757 | Median follow up - Unvaccinated - 66.0 (66.0-66.0), Vaccinated - 63.0 (52.0-65.0) | -- | -- | -- |
| IQR: Interquartile range, PV: Partial vaccination, FV: Full vaccination, UV: Unvaccinated, NR: Not reported | | | | | | | | | | |
