## Supplementary material for "Postvaccination SARS-CoV-2 infection among healthcare workers – A Systematic Review and meta-analysis": Table 1

**Table 1 – Study Characteristics and Patient Details**

| Author | Design | Vaccine Type | Total Patients Enrolled | Total COVID-19 Positive Patients | Age – Years [SD] (Range) | Sex (M/F) | Patients | | | COVID-19 Positive | | |
| --- | --- | --- | --- | --- | --- | --- | --- | --- | --- | --- | --- | --- |
|  |  |  |  |  |  |  | PV | FV | UV | PV | FV | UV |
| Amit, 2021 | Retrospective, Single Center, December 2020 - January 2021, Israel. | Pfizer-BioNTech COVID-19 vaccine (BNT162B2) | 4,081 | 22 | 45.3 (9.85) [31-61] | 8/ 14 (positives) | 4,081 | -- | -- | 22/4081 | -- | -- |
| Azamgarhi, 2021 | Retrospective, Single Center, United Kingdom. | Pfizer-BioNTech COVID-19 vaccine (BNT162B2) | 2,235 | 49 | 584 (16–34), 1234 (35–54), 417 (>55) | 722/1513 (total) | 1,409 | -- | 826 | 23/1409 | -- | 26/826 |
| Bouton, 2021 | Prospective, Single Center, December 9, 2020 and February 23, 2021, USA | Pfizer-BioNTech COVID-19 Vaccine (BNT162B2) and Moderna Covid Vaccine (mRNA-1273) | 10,590 | 425 | 40 [13] | 18/78 (positives) | 7,109 | 5,913 | 3,481 | 96/7109 | 17/5913 | 329/3481 |
| Tang, 2021 | Prospective, Single Center, December 17, 2020 - March 20, 2021, USA | Pfizer-BioNTech COVID-19 vaccine (BNT162B2) | 5,217 | 236 | -- | 1038/2014 (vaccinated) | 3,052 | 2,776 | 2165 | 41/3052 | 10/2776 | 185/2165 |
| Cucunawangsiha, 2021 | Retrospective, Single, Center, Indonesia | 2021, CoronaVac (Sinovac Biotech) | 1,040 | 13 | 32.5 [5.9] | 5/8/2021 (positives) | -- | 1,040 | -- | -- | 13/1040 | -- |
| Hall, 2021 | Prospective, Multi-center, December 8, 2020 - February 5, 2021, USA | Pfizer-BioNTech COVID-19 vaccine (BNT162B2) and Oxford-AstraZeneca COVID-19 Vaccine (ChAdOx1 nCoV-19 adenoviral [AZD1222]) | 23,324 | 80 | 46.1 (36 - 54.1) | 3632/19692 (total) | 20,641 | 1,607 | 2683 | 71/20641 | 9/1607 | 977/2683 |
| Issac, 2021 | Prospective, NR, Single Center, January 27, 2021 - July 15, 2021, India | ChAdOx1 nCoV-19 | 324 | 41 | 34.09 [9.43] | 52/272 (vaccinated) | -- | 243 | 80 | -- | 16/243 | 35/80 |
| Jacobson, 2021 | Retrospective, December 18, 2020 - April 02, 2021, USA | Pfizer-BioNTech COVID-19 Vaccine (BNT162B2) and Moderna Covid Vaccine (mRNA-1273) | 30,000 | 189 | 40.8 (11.2) [Mean], 38 (32–48) [Median] | 64/125 (positive) | 23,090 | 22,271 | 6,910 | 150/23090 | 39/22271 | 471/6910 |
| Maroof, 2021 | Prospective, Feburay 2021 - June 2021, Multicenter, Pakistan | Sino-pharm Vaccine | 39,512 | 124 | 38.8 ± 11 | 86/38 (positives) | -- | 39,512 | -- | -- | 124/39512 | -- |
| Mathema, 2021 | Retrospective, December 2020 to April 2021, Multicenter, USA | Pfizer-BioNTech COVID-19 Vaccine (BNT162B2) and Moderna Covid Vaccine (mRNA-1273) | 37,500 | 138 | 45 (19-77) [Median] [N=121] (postives) | 31/90 [N=121] (positives) | 23,697 | 22,458 | -- | 105/138 | 33/22458 | -- |
| North, 2021 | Prospective, cohort, December 30, 2020 - April 2, 2021, Multicenter, USA | Pfizer-BioNTech COVID-19 Vaccine (BNT162B2) and Moderna Covid Vaccine (mRNA-1273) | 2,247 | 19 | 37 (30-50) | 476/1760 (total) | 2,036 | 1,923 | 593 | 10/2036 | 3/1923 | 6/593 |
| Pandurangaiah, 2021 | Retrospective, Feburary 2021 - May 07, 2021, Single center, India | Covishield/Covaxin | 182 | 6 | -- | 47/135 (total) | 18 | 164 | -- | 1/18 | 5/164 | -- |
| Sabnis, 2021 | Prospective, First dose of COVID-19 vaccine till June 10, 2021, Single center, India | Covishield (SII-ChAdOx1 nCoV-19) | 461 | -- | 34.18 (22-77) | NR | -- | 461 | -- | -- | 86/461 | -- |
| Vaishya, 2021 | Retrospective, observational, cohort, January 16 2021 - June 15 2021, Multicenter, India | AstraZeneca Covid Vaccine (ChAdOx1-S/nCoV-19 [recombinant]) (Covishield 26,375, Covaxin 1,967) | 28,342 | 1,438 | 33.04 (18-80) | 14980/13362 (total) | 5,125 | 23,217 | -- | 269/5125 (within 2 weeks 46, after 2 weeks 223) | 1169/23217 ((within 2 weeks 69, after 2 weeks 1100) | -- |
| Keehner, 2021 | Retrospective, December 16, 2020 - February 9, 2021, Multicenter, USA | Pfizer-BioNTech COVID-19 Vaccine (BNT162B2) and Moderna Covid Vaccine (mRNA-1273) | 36,659 | 379 | -- | -- | 36,659 | 28,184 | -- | 342/36659 | 37/28184 | -- |
| Tyagi, 2021 | Retrospective, January 16, 2021 - till date, Single center, India | 85 (Covishield), 28 (Covaxin) 113 (vaccinated) | 123 | 19 | 42 (22-70) | 75/48 (total) | 6 | 107 | 10 | 1/6 | 18/107 | -- |
| Sharma, 2021 | Cross-sectional study (May 31, 2021 - June 6, 2021), January 2021 - March 2021 (Subjects Data), Single center, India | 158 (Covishield), 168 (Covaxin) | 326 | 65 | 29.1 (85.9) | 212/114 | 46 | 280 | -- | 12/46 | 53/280 | -- |
| Angel, 2021 | Single-center, retrospective cohort study, December 20, 2020, and February 25, 2021, Israel | Pfizer-BioNTech BNT162b2 | 6710 | 270 | 44.3 [12.5] | 2245/4465 | 5953 | 5517 | 757 | 127/5761 | 27/5372 | 116/757 |
| PV: Partial vaccination, FV: Full vaccination, UV: Unvaccinated | | | | | | | | | | | | |
