## Supplementary Material for "Postvaccination SARS-CoV-2 infection among healthcare workers – A Systematic Review and meta-analysis"

SUPPLEMENTARY MATERIALS

APPENDIX-1: Literature search strategy

APPENDIX-2: MOOSE Checklist

APPENDIX-3: PRISMA Flow Diagram

Supplementary Table 1: Newcastle-Ottawa Scale - Study quality assessment

Supplementary Figures 2: Forest Plot, COVID-19 infections in unvaccinated HCWs

Supplementary Figures 3: Forest Plot, vaccinated HCWs hospitalized for COVID-19 infection

Supplementary Figures 4: Forest Plot, vaccinated HCWs admitted to ICU for COVID-19 infection

Supplementary Figures 5: Forest Plot, vaccinated HCWs died from COVID-19 infection

Supplementary Figures 6 (a-c): Funnel Plot, Publication Bias

APPENDIX-1: Literature search strategy

| Literature Database | Search items | Items found |
| --- | --- | --- |
| ***Embase via Ovid (1974+)*** | ('covid-19'/exp OR 'covid-19' OR 'coronavirus disease') AND ('health care worker'/exp OR 'health care worker') AND ('vaccine'/exp OR 'vaccine') | 475 |
| ***MEDLINE via Ovid (1946+)*** | "covid-19" OR "coronavirus disease" AND "health care worker" AND "vaccine" | 25 |
| ***Scopus via Elsevier (1970+)*** | TITLE-ABS-KEY ( "covid-19"  AND  "health care workers"  AND  "vaccine" ) | 186 |
| ***Web of Science Core Collection via Clarivate Analytics (1975+)*** | "covid-19"  OR  "coronavirus disease"  AND  "health care workers" AND “vaccine” | 428 |

APPENDIX-2: MOOSE Checklist

| Item No | Recommendation | Reported on Page No |
| --- | --- | --- |
| Reporting of background should include | | |
| 1 | Problem definition | 4-5 |
| 2 | Hypothesis statement | 5 |
| 3 | Description of study outcome(s) | 5 |
| 4 | Type of exposure or intervention used | 5 |
| 5 | Type of study designs used | 5 |
| 6 | Study population | 6 |
| Reporting of search strategy should include | | |
| 7 | Qualifications of searchers (eg, librarians and investigators) | 5 |
| 8 | Search strategy, including time period included in the synthesis and key words | 5 |
| 9 | Effort to include all available studies, including contact with authors | 6 |
| 10 | Databases and registries searched | 5 |
| 11 | Search software used, name and version, including special features used (eg, explosion) | 5 |
| 12 | Use of hand searching (eg, reference lists of obtained articles) | -NA- |
| 13 | List of citations located and those excluded, including justification | 8-9,  Suppl Appendix 3 |
| 14 | Method of addressing articles published in languages other than English | -NA- |
| 15 | Method of handling abstracts and unpublished studies | 6 |
| 16 | Description of any contact with authors | -NA- |
| Reporting of methods should include | | |
| 17 | Description of relevance or appropriateness of studies assembled for assessing the hypothesis to be tested | 5-6 |
| 18 | Rationale for the selection and coding of data (eg, sound clinical principles or convenience) | 6 |
| 19 | Documentation of how data were classified and coded (eg, multiple raters, blinding and interrater reliability) | 6 |
| 20 | Assessment of confounding (eg, comparability of cases and controls in studies where appropriate) | 6 |
| 21 | Assessment of study quality, including blinding of quality assessors, stratification or regression on possible predictors of study results | 7 |
| 22 | Assessment of heterogeneity | 8 |
| 23 | Description of statistical methods (eg, complete description of fixed or random effects models, justification of whether the chosen models account for predictors of study results, dose-response models, or cumulative meta-analysis) in sufficient detail to be replicated | 8 |
| 24 | Provision of appropriate tables and graphics | Tables 1-2, Figs 1-3 |
| Reporting of results should include | | |
| 25 | Graphic summarizing individual study estimates and overall estimate | Fig 1-3 Suppl Fig 1-5 |
| 26 | Table giving descriptive information for each study included | Table 1-2 |
| 27 | Results of sensitivity testing (eg, subgroup analysis) | 12-13 |
| 28 | Indication of statistical uncertainty of findings | 12-13 |

APPENDIX-3: PRISMA Flow Diagram

Records excluded
(n = 161)

Records screened
(n = 244)

Records after duplicates removed
(n = 477)

### Identification

### Eligibility

### Included

### Screening

Records identified through database searching
(n = 1114)

Full-text articles excluded, with reasons
(n = 65)

(Total vaccinated HCWs not reported, letter to the editors, Systematic Reviews)

S

Full-text articles assessed for eligibility
(n = 83)

Studies included in quantitative synthesis (meta-analysis)
(n = 18)

Supplementary Table 1: Newcastle-Ottawa Scale - Study quality assessment

| **STUDY** | **SELECTION** | | | | **COMPARABILITY** | **OUTCOME** | | | **SCORE** | **QUALITY** |
| --- | --- | --- | --- | --- | --- | --- | --- | --- | --- | --- |
|  | **Representativeness of the average adult in community** | **Cohort size** | **Information on clinical outcomes** | **Outcome not present at start** | **Factors comparable between the groups** | **Adequate clinical assessment** | **Follow up time** | **Adequacy of follow-up** | **MAX=8** | **HIGH>6, MEDIUM 4 to 6, LOW <4** |
|  | **Population based: 1; Multi-center: 0.5; Single-center: 0** | **>40 patients: 1; 39 to 20: 0.5; <20: 0** | **Information with clarity: 1; Information derived from percentage value: 0.5; Unclear: 0** | **not present: 1; present: 0** | **yes: 1; no: 0** | **yes: 1; no: 0** | **yes: 1; not mentioned: 0** | **All patients followed up: 1; >50% followed up: 0.5; <50% followed up OR not mentioned: 0** |  |  |
| Amit, 2021 | 0 | 1 | 1 | 1 | 1 | 1 | 1 | 1 | 7 | HIGH |
| Azamgarhi, 2021 | 0 | 1 | 1 | 1 | 1 | 1 | 1 | 1 | 7 | HIGH |
| Bouton, 2021 | 0 | 1 | 1 | 1 | 1 | 1 | 1 | 1 | 7 | HIGH |
| Tang, 2021 | 0 | 1 | 1 | 1 | 1 | 1 | 1 | 1 | 7 | HIGH |
| Cucunawangsiha, 2021 | 0 | 1 | 1 | 1 | 1 | 1 | 1 | 1 | 7 | HIGH |
| Hall, 2021 | 0.5 | 1 | 1 | 1 | 1 | 1 | 1 | 1 | 6.5 | HIGH |
| Issac, 2021 | 0 | 1 | 1 | 1 | 1 | 1 | 1 | 1 | 7 | HIGH |
| Jacobson, 2021 | 0 | 1 | 1 | 1 | 1 | 1 | 1 | 1 | 7 | HIGH |
| Maroof, 2021 | 0.5 | 1 | 1 | 1 | 1 | 1 | 1 | 1 | 6.5 | HIGH |
| Mathema, 2021 | 0.5 | 1 | 1 | 1 | 1 | 1 | 1 | 1 | 6.5 | HIGH |
| North, 2021 | 0.5 | 1 | 1 | 1 | 1 | 1 | 1 | 1 | 6.5 | HIGH |
| Pandurangaiah, 2021 | 0 | 1 | 1 | 1 | 1 | 1 | 1 | 1 | 7 | HIGH |
| Sabnis, 2021 | 0 | 1 | 1 | 1 | 1 | 1 | 1 | 1 | 7 | HIGH |
| Vaishya, 2021 | 0.5 | 1 | 1 | 1 | 1 | 1 | 1 | 1 | 6.5 | HIGH |
| Keehner, 2021 | 0.5 | 1 | 1 | 1 | 1 | 1 | 1 | 1 | 6.5 | HIGH |
| Tyagi, 2021 | 0 | 1 | 1 | 1 | 1 | 1 | 1 | 1 | 7 | HIGH |
| Sharma, 2021 | 0 | 1 | 1 | 1 | 1 | 1 | 1 | 1 | 7 | HIGH |

Supplementary Figure 2: Forest Plot, COVID-19 infections in unvaccinated HCWs

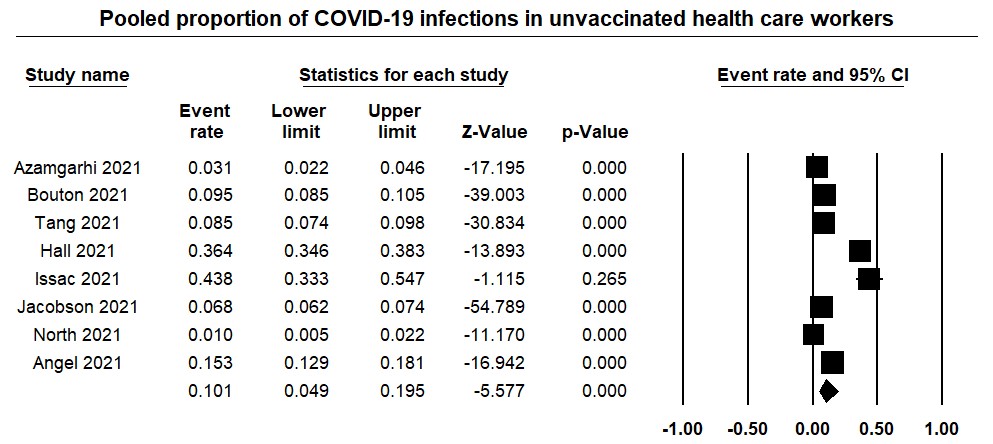

Supplementary Figure 3: Forest Plot, vaccinated HCWs hospitalized for COVID-19 infection

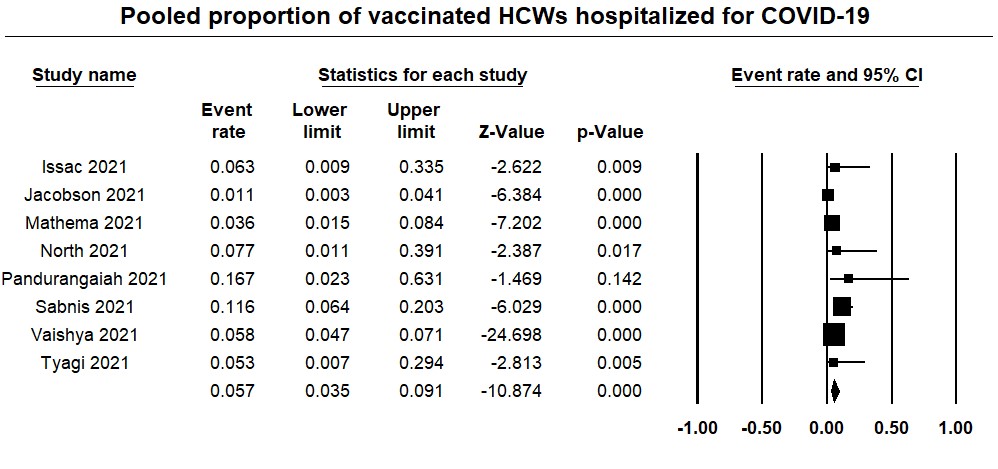

Supplementary Figure 4: Forest Plot, vaccinated HCWs admitted to ICU for COVID-19 infection

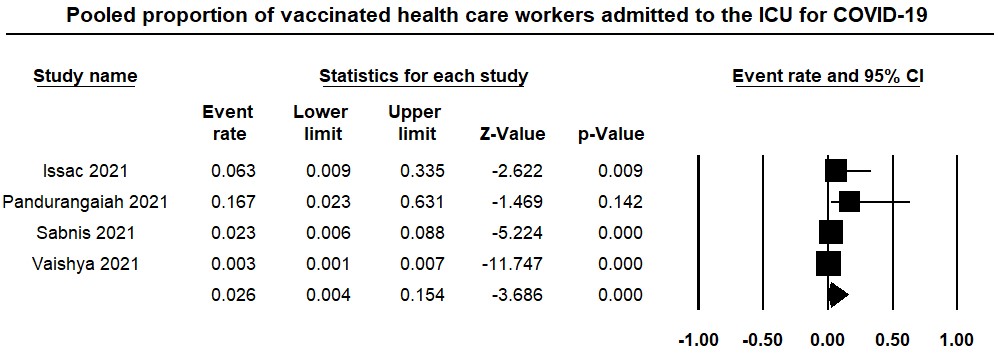

Supplementary Figure 5: Forest Plot, vaccinated HCWs died from COVID-19 infection

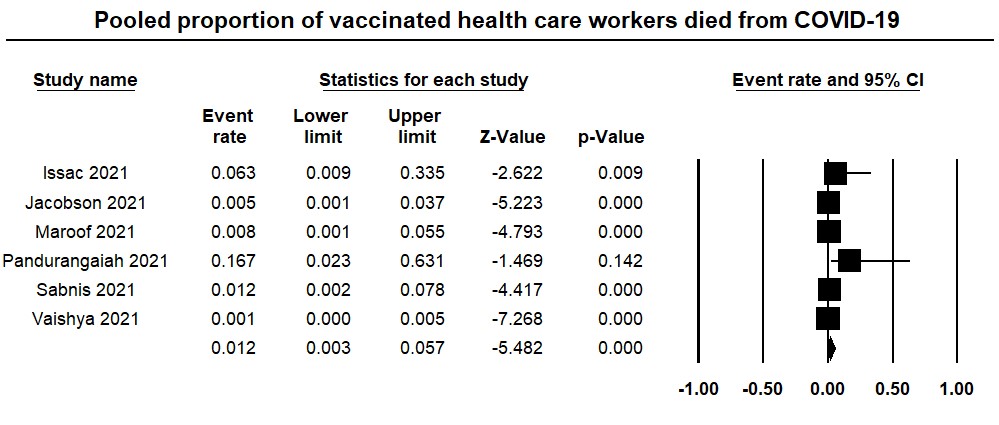

Supplementary Figures 6 (a-c): Fu
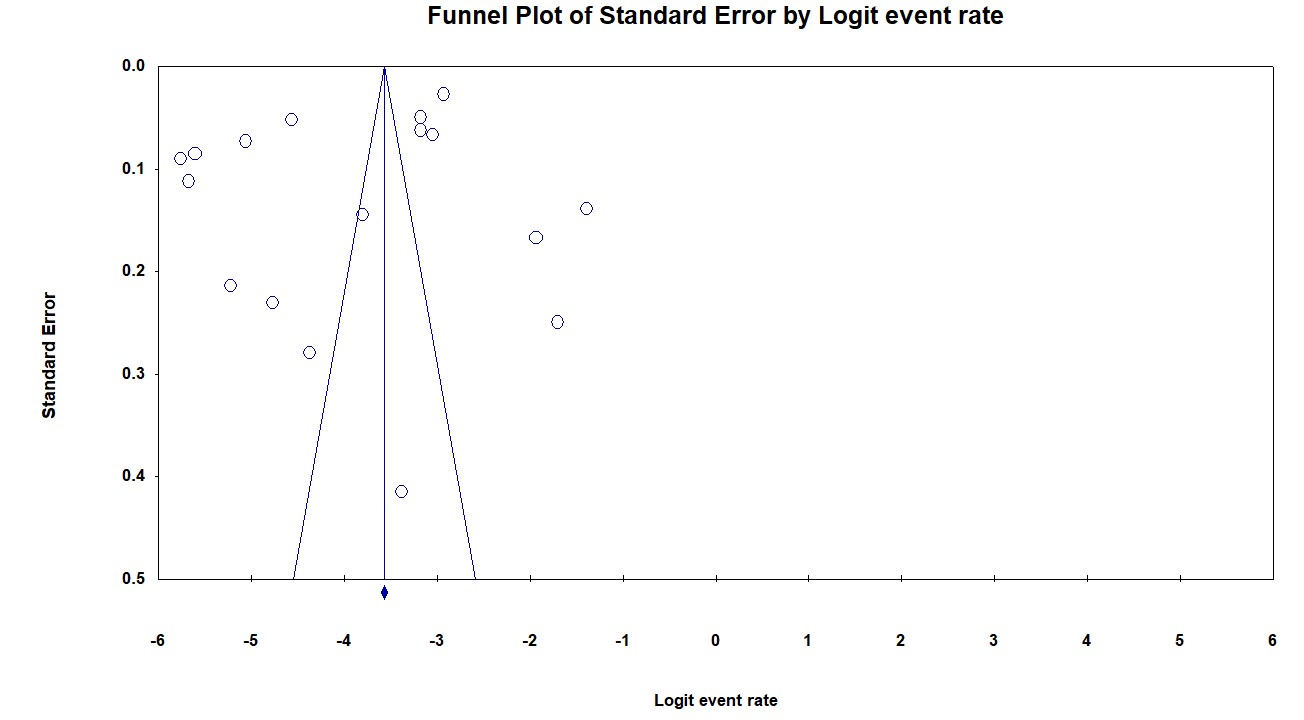
nnel Plot, Publication Bias

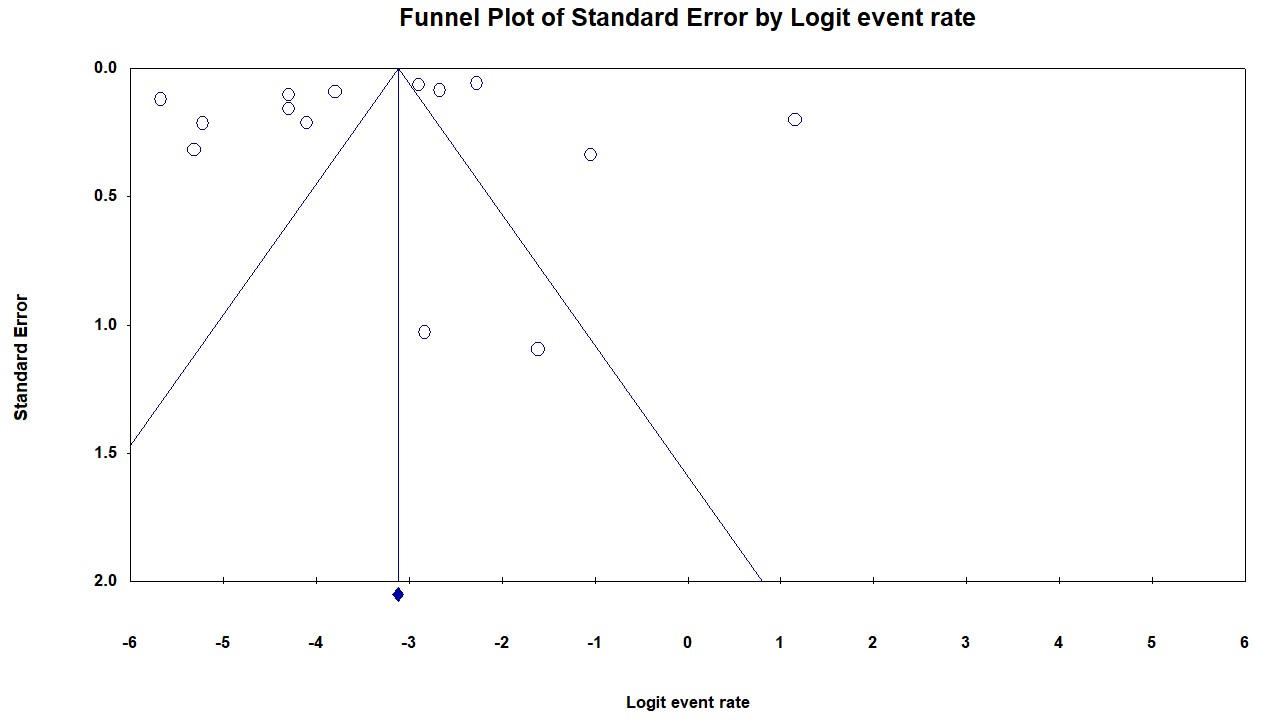

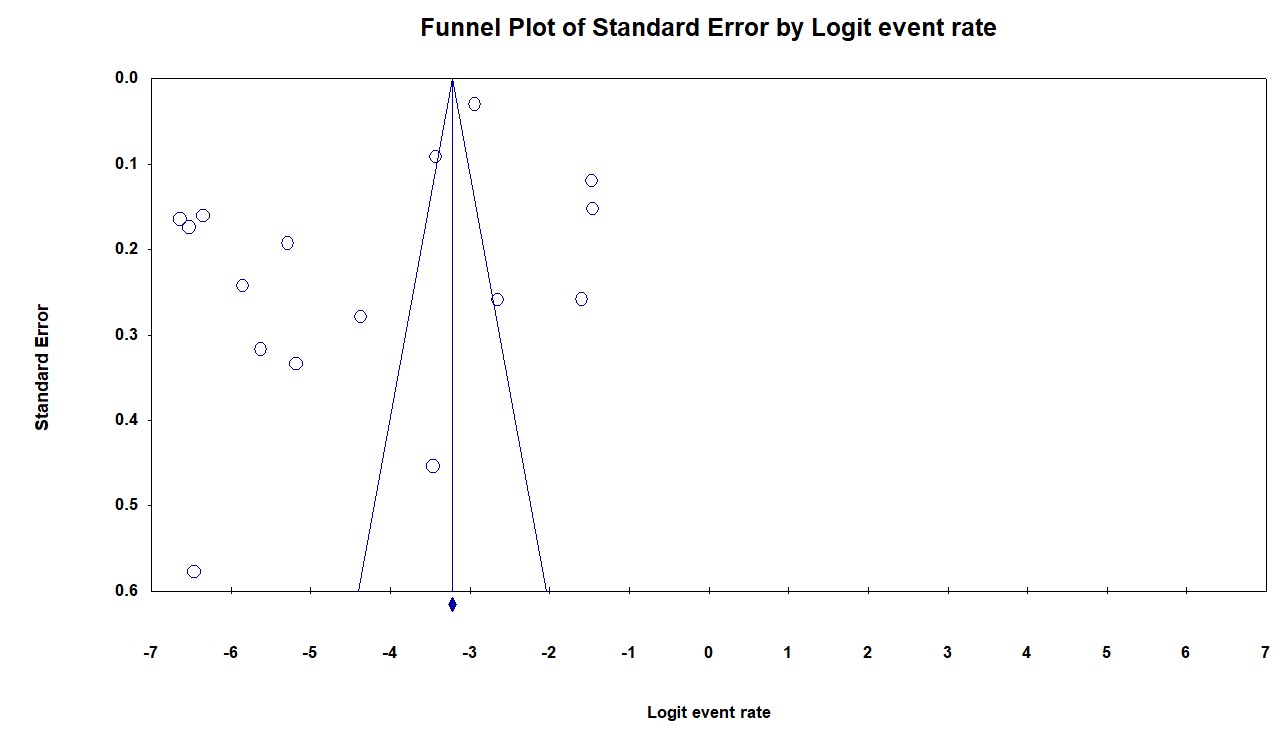
